# Emergent visual creativity in frontotemporal dementia is associated with dorsomedial visual cortex enhancement

**DOI:** 10.1101/2022.05.31.22275081

**Authors:** Adit Friedberg, Lorenzo Pasquini, Ryan Diggs, Erika A. Glaubitz, Lucia Lopez, Ignacio Illán-Gala, Leonardo Iaccarino, Renaud La Joie, Nidhi Mundada, Jesse Brown, Isabel Elaine Allen, Katherine P. Rankin, Luke W. Bonham, Jennifer S. Yokoyama, Eliana M. Ramos, Daniel H. Geschwind, Salvatore Spina, Lea T. Grinberg, Zachary A. Miller, Joel H. Kramer, Howard Rosen, Maria Luisa Gorno-Tempini, Gil Rabinovici, William W. Seeley, Bruce L. Miller

## Abstract

**IMPORTANCE:** The neurological substrates of visual creativity are unknown. We demonstrate the role of dorsomedial visual cortex in emergence of visual artistic creativity (VAC) in the setting of dementia. Our findings illuminate neural substrates of human creativity and suggest that hyperactivation of specific brain areas may manifest as enhanced cognitive or behavioral capacities.

**OBJECTIVE:** To determine the anatomical and physiological underpinnings of VAC in dementia.

**DESIGN, SETTING, AND PARTICIPANTS:** As part of a prospective, longitudinal cohort study focused on frontotemporal dementia (FTD), 734 patients met research criteria for an FTD spectrum disorder between 2002 and 2019. Of these, seventeen showed emergence of visual artistic creativity (VAC-FTD). Two control groups (n = 51 each) were matched to VAC-FTD based on demographic and clinical parameters: (1) Not Visually Artistic FTD (NVA-FTD) and (2) Healthy Controls (HC).

**MAIN OUTCOMES AND MEASURES:** Clinical, neuropsychological, genetic and neuroimaging data were analyzed to characterize VAC-FTD and compare VAC-FTD to control groups.

**RESULTS:** Emergence of VAC occurred around the time of onset of symptoms, and was disproportionately seen in patients with temporal lobe predominant degeneration (n = 8/17). Atrophy network mapping identified a dorsomedial occipital region whose activity inversely correlated, in healthy brains, with activity in the patient-specific atrophy patterns in VAC-FTD (n = 17/17) and NVA-FTD (n = 45/51). Structural covariance analysis revealed that volume of this dorsal occipital region was strongly correlated, in VAC-FTD, but not in NVA-FTD or HC, with a volume in the primary motor cortex corresponding to the right hand representation. One patient, who underwent fluorodeoxyglucose positron emission tomography before and after VAC onset, showed increasing glucose metabolism in the dorsal occipital region over the interval when creativity emerged.

**CONCLUSIONS AND RELEVANCE:** FTD lesion-induced intensification of dorsal visual association cortex structure and function predisposes to emergence of VAC in certain environmental or genetic conditions. Paradoxical gains of function are early manifestations of neurodegenerative disease, and this study delineates a specific brain region associated with the emergence of VAC.

## Introduction

Creativity, the ability to generate work that is both novel and valuable^1^, is pivotal to the development of human culture, as it enables transformative problem solving, technological progress, and artistic expression. Visual artistic creativity (VAC) is defined as the production of novel and aesthetically pleasing visual forms, and is a process that depends heavily on visual mental imagery^2^. VAC is unique to and ubiquitous in human societies^3^, and insights into VAC have the potential to Illuminate the neural underpinnings of creativity more broadly^4-7^. Frontotemporal dementia (FTD) represents a group of neurodegenerative disorders characterized by progressive deterioration of behavior and/or language, most often associated with focal frontotemporal lobar degeneration (FTLD) pathology affecting the frontal, insular, and temporal cortex. Emergence of novel visual artistic skills has been described in FTD, particularly in the primary progressive aphasia (PPA) variants ^8-11^, which result from degeneration of the left frontal and anterior temporal lobes. Moreover, patients with focal brain lesions caused by stroke and traumatic brain injury have been reported to develop new visual artistic skills^12-14^, most often after injury to brain regions affected in FTD. Therefore, patients with anterior brain lesions provide a rare window into the neural network building blocks of VAC. Previously, we speculated that selective degeneration of the frontal and anterior temporal lobes, within the language-dominant hemisphere, led to decreased inhibition of posterior visual-spatial systems involved in visual perception and association, thereby enhancing artistic interest^11^. This hypothesis has not been systematically examined, and the underlying neural mechanisms for VAC in the setting of brain injury remain uncertain.

We describe the clinical, neuropsychological, neuropathological, and genetic features of seventeen patients with FTD, drawn from a cohort of 734 patients assessed over seventeen years, who reported emergence of VAC. We then probe the neural substrates of this phenomenon using atrophy network mapping and structural covariance analyses, applied to patients with FTD with and without VAC and controls.

## Methods

### Participants

Patients were evaluated at the University of California, San Francisco, Memory and Aging Center as part of a prospective, longitudinal cohort study focused on FTD spectrum disorders. Visits occurred between January 2002 and May 2019. Patients underwent a standardized clinical, neuropsychological, and neuroimaging ^15^ evaluations. Genotyping for autosomal dominant pathogenic genetic variants in *MAPT, GRN* and *C9orf72* was performed as described^16^. Inclusion criteria included a clinical diagnosis of behavioral variant FTD (bvFTD), non-fluent variant PPA (nfvPPA), semantic variant PPA (svPPA), progressive supranuclear palsy – Richardson syndrome (PSP-RS) corticobasal syndrome (CBS) or amyotrophic lateral sclerosis (ALS), diagnosed according to prevaling clinical research criteria at the time of assessment^17-24^. These criteria yielded a pool of 734 patients, from which, 45 were excluded due to lack of available clinical records. For the remaining 689 patients, retrospective chart review was performed by a single investigator (A.F., a behavioral neurologist) to identify patients who met the following criteria for emergent VAC: (1) emergence of novel visual artistic skills, (2) a change in the style of visual art produced, or (3) a substantial increase in quantity of visual art generated. Demographic features and description of visual artistic behavior were recorded, and all available artwork was collected. To evaluate the neural signature unique to patients with emergence of VAC in the setting of FTD, two control groups were assembled: (1) Not Visually Artistic FTD (NVA-FTD) and (2) Healthy Controls (HC). NVA-FTD included patients with FTD spectrum disorders for whom no change in visual artistic behavior was reported. NVA-FTD patients were matched to the VAC-FTD group by clinical diagnosis, disease stage (assesessed using the Clinical Dementia Rating Scale sum of boxes score (CDR-SB))^25^, age, sex, handedness, and years of education. A matching process accounting for multiple variables was required, and we matched three controls to each VAC-FTD participant. The HC group was matched for age, sex, handedness and years of education (Table 1). Between-group differences in clinical and neuropsychological characteristics were assessed using parametric or nonparmteric tests as appropriate. Test statistics were considered significant at *P* < 0.05 (two-tailed, eMethods 1).

**Table 1:**
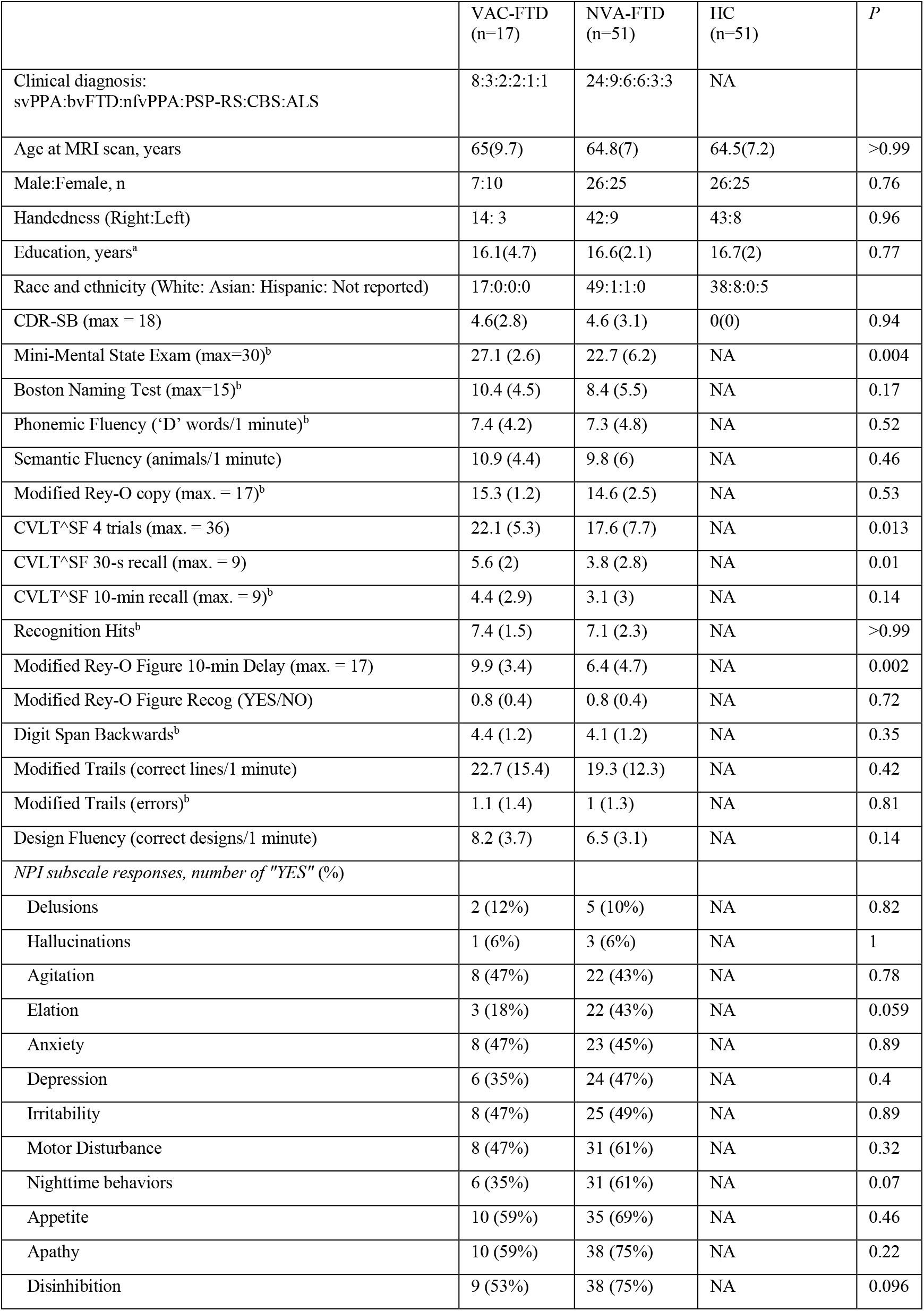
Participants’ demographic and neuropsychological features. Means with standard deviations in parentheses reported unless noted otherwise. ^a^Kruskal-Wallis test was used, ^b^Mann-Whitney test was used All cognitive measures were available for at least 80% of participants in each group. NPI measures were available for all participants. Abbreviations: VAC-FTD, patients with a frontotemporal dementia spectrum disease and emergence of visual artistic creativity; NVA -FTD, patients with a frontotemporal dementia spectrum disease without emergence of visual artistic creativity; HC, healthy controls; svPPA, semantic variant of primary progressive aphasia; bvFTD, behavioral variant of frontotempotral dementia; nfvPPA nonfluent variant of primary progressive aphasia; PSP-RS progressive supranuclear palsy - Richardson syndrome ; CBS, corticobasal syndrome;ALS amyotrophic lateral sclerosis; MRI, magnetic resonance image; CDR-SB, Clinical Dementia Rating scale sum of boxes; MMSE, Mini-Mental State Examination; CVLT^SF, California Verbal Learning Test Short Form; NPI, Neuropsychiatric Inventory; NA, not applicable.

### Structural MRI acquisition and preprocessing

Over the ascertainment period, MR images were acquired with four different scanners, using several image acquisition protocols (eTable 1, eMethods 2). Magnetic field strength was 1.5, 3.0, or 4.0 Tesla. Structural images were segmented into gray matter, white matter, and cerebrospinal fluid and normalized to MNI space using SPM12 (http://www.fil.ion.ucl.ac.uk/spm/software/spm12/). Gray matter images were modulated by dividing the tissue probability values by the Jacobian of the warp field and smoothed with an isotropic Gaussian kernel with a full width at half maximum of 8 mm^26^.

### Individual atrophy (W-score) maps

To generate participant-specific atrophy maps, the smoothed gray matter images were transformed into W-score maps (see eMethods 3). W-score maps are voxel-wise statistical maps that reflect levels of atrophy for each individual after adjustment for relevant covariates^27,28^. The W-score model generated for this study was based on 397 healthy controls and included age at MRI, sex, years of education, handedness, scanner type, and total intracranial volume as covariates.

Individual W-score maps were thresholded and binarized to capture each patient’s 1% most atrophied voxels. This procedure enabled us to represent patients’ focal neurodegeneration in a balanced manner across subjects of varying overall atrophy severity and extent. To ensure robustness of findings across a range of atrophy thresholds, we repeated all analyses with more (highest 0.5%) and less (highest 2%, 5%) stringent thresholds.

### Comparing W-score maps in VAC-FTD versus NVA-FTD

We compared the unthresholded W-score maps between groups (VAC-FTD versus NVA-FTD) at every voxel using a two-sample t-test. Mini-Mental State Exam (MMSE)^29^ score was included as a nuisance covariate. Significant clusters were identified, across the whole brain, using a t-threshold corresponding to *P* < 0.001 uncorrected for multiple comparisons.

### Atrophy Network Mapping

Next, we sought to determine how the brain areas atrophied in patients are functionally connected, in the healthy brain, to other brain regions. First we derived an ‘atrophy network map’ for each patient, seeded by each patient’s binarized single-subject atrophy map (top 1% most atrophied voxels)^30-33^. Using this seed region, we turned to task-free fMRI data from a cohort of 175 cognitively healthy individuals, matched to the VAC-FTD by age, sex, handedness, and years of education (eTable 2). In these subjects, we computed the average blood oxygen level-dependent (BOLD) signal time series for all voxels within the patient-derived atrophy seed region and correlated these mean time series with the time series of every other voxel (eMethods 4). Resulting r-values were converted to a normal distribution using Fisher’s r-to-z transform and entered into a single, group-level, voxel-wise one-sample t-test. The resulting maps constituted the unthresholded atrophy network t-maps. Positive and negative functional correlations were thresholded at t ≥ |7|, corresponding to *P* < 10^−10^, to create a binarized map of connected regions, in keeping with prior approaches^34^. To ensure that results were not dependent on this threshold, we repeated our analysis with t-value thresholds of 6 (*P*<10^−7^) and 8 (*P*< 10^−12^). Finally, the binarized atrophy network maps were overlaid to generate frequency maps for each patient group, representing the proportion of patients in that group whose lesions were functionally connected to each voxel in the healthy brain (eFigure 1). We also compared the unthresholded atrophy network t-maps between the VAC-FTD and NVA-FTD groups, voxel-wise, using a two-sample t-test. Significant clusters were identified, across the whole brain, using a t-threshold corresponding to *P* < 0.001, uncorrected for multiple comparisons.

### Structural covariance analysis

Atrophy network mapping results were used as the basis for investigating differences in inter-regional structural correlations between VAC-FTD and NVA-FTD^35,36^. To that end, a voxel-wise interaction model was implemented in SPM12 using the W-score maps of the VAC-FTD and NVA-FTD groups and adding a term for the individual mean W-score of the region of interest (ROI) as a covariate. An interaction term between group membership (VAC-FTD/NVA-FTD) and individual mean W-score of the ROI was included, and a statistical contrast was set to elicit group differences in covariance between the individual mean W-score of the ROI and other brain regions, with MMSE included as a confounding covariate. A second interaction model examined group differences in structural covariance to the ROI in VAC-FTD versus HC. In this model, MMSE was omitted because it is highly confounded with group. A statistical threshold of *P* < 0.001 (whole brain, uncorrected) with a minimum cluster extent of 100 voxels and cluster level threshold of *P* < 0.05 (uncorrected) was applied for both interaction models.

### FDG PET

18F-labeled fluorodeoxyglucose positron emission tomography (FDG-PET) images were acquired at Lawrence Berkeley National Laboratory using a Siemens ECAT EXACT HR PET scanner in 3-dimensional acquisition mode. PET frames were preprocessed using standard methods and standardized uptake value ratio (SUVR) maps were created (eMethods 5). Based on SUVR maps of 71 healthy controls, a voxel-wise FDG-PET W-score model was generated including age, sex, handedness, and education as covariates. For a single patient who had FDG PET scanning 14 months apart, before and after emergence of VAC, individual voxel-wise FDG-PET W-score maps were generated. For these two maps, mean regional SUVR W-scores were extracted from Brainnetome atlas parcels^37^ (eMethods 6).

## Results

### Clinical and demographic characteristics

Among the 689 patients with FTD whose clinical records were reviewed, seventeen met the operational definition of emergent VAC, with a resulting prevalence of 2.5%. Eight of seventeen patients had de novo emergence of VAC, seven showed some past interest in either visual or non-visual art, and two were artists who experienced substantial change in artistic style. The most frequently associated FTD clinical syndrome was semantic variant primary progressive aphasia (svPPA), accounting for nearly half of the cases and for 6.7% of all patients with svPPA who were screened (Figure 1, eTable 3). No pathogenic variants in *C9orf72, GRN*, or *MAPT* were found in the 15/17 VAC-FTD patients who underwent genetic testing. Neuropathological diagnoses available for 6/17 VAC-FTD patients revealed diverse underlying FTLD subtypes (eTable 4).

**Figure 1.**
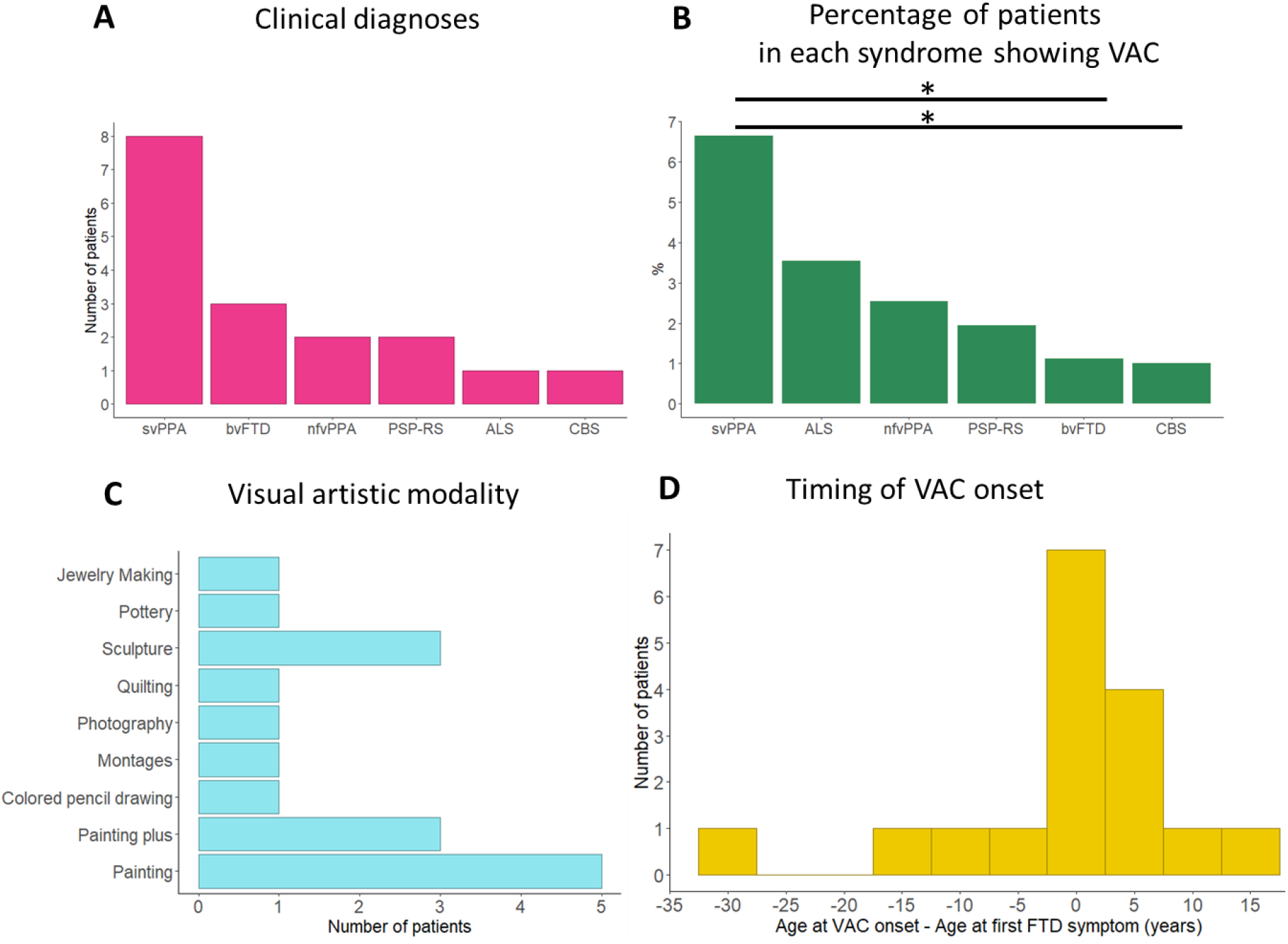
Clinical characteristics and artistic expression in patients with frontotemporal dementia who reported emergence of visual artistic creativity (VAC-FTD, n = 17). **A**, SvPPA was the most common clinical syndrome in the VAC-FTD group. **B**, SvPPA was also the FTD syndrome with the highest percentage of patients with VAC relative to the overall syndrome-based cohort, significantly higher than in bvFTD and CBS (Fisher’s exact test, *P* < 0.05). **C**, Painting was the most common modality of visual artistic expression. “ Painting plus” indicates that painting, the primary modality was accompanied by other forms of visual art: sculpture (n = 1), photography (n = 1), jewelry making, and glass painting (n = 1). **D**, Most patients showed emergence of visual artistic creativity in close temporal proximity to FTD symptom onset. Abbreviations: svPPA, semantic variant of primary progressive aphasia; bvFTD, behavioral variant of frontotempotral dementia; nfvPPA, nonfluent variant of primary progressive aphasia; PSP-RS, progressive supranuclear palsy with Richardson syndrome; CBS, corticobasal syndrome; ALS, amyotrophic lateral sclerosis; VAC, visual artistic creativity.

Emergence of VAC occurred early in the FTD disease course. Most patients experienced the change at, or shortly after, the time of FTD symptom onset, but 4/17 patients showed emergent VAC before their FTD symptoms appeared (Figure 1). As mandated by the study design VAC-FTD and NVA-FTD showed no significant differences in demographic or clinical variables. MMSE score was higher in VAC-FTD than NVA-FTD reflecting better preserved memory in this group (Table 1). Consequently, MMSE was used as nuisance covariate in neuroimaging analyses. Prevalence of neuropsychiatric symptoms measured by the Neuropsychiatric Inventory was similar in the two groups^38^.

### Visual art

Visual art collected from 11/17 patients included painting, quilting, jewelry making, sculpture, pottery, and montage making (eTable 3). Bright colors were common, and the art rarely focused on human faces. In some, there was evidence for loss of semantic knowledge. For example, two patients with svPPA generated animal sculptures lacking the features of a species, producing generic or prototypical representations of an “ animal”. When humans and animals were depicted, facial expressions were often bizarre and did not convey natural emotions, as has been previously described (Figure 2) ^39,40^.

**Figure 2.**
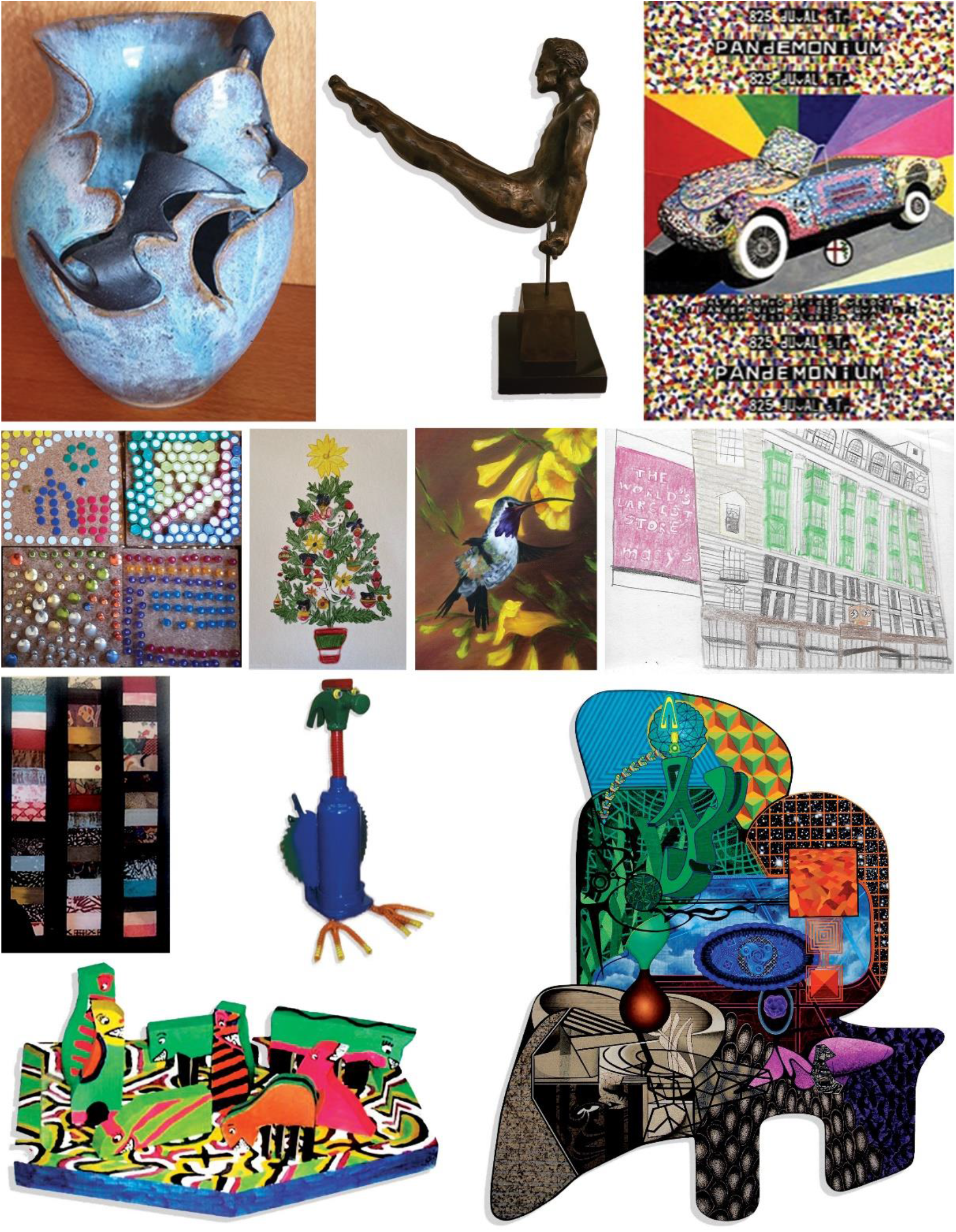
Examples of the visual artworks. Each piece was selected to represent the style of a single patient in the VAC-FTD group (examples were available for 11/17).

### Structural MRI

Patients in the VAC-FTD and NVA-FTD groups showed typical group-level atrophy maps highlighting neurodegeneration in the anterior temporal lobes (left greater than right), amygdalae, striatum, and left insula (eFigure 2,3). Statistical W-score map and gray matter map comparisons between VAC-FTD and NVA-FTD revealed no group differences.

### Atrophy Network Mapping

Individual atrophy network maps were thresholded at t ≥ |7|, binarized, and overlayed to generate a group level atrophy network frequency map for the VAC-FTD and NVA-FTD groups. Group level atrophy network frequency maps identified a bilateral dorsomedial occipital region anticorrelated, in the healthy brain, to the atrophy patterns of 17/17 VAC-FTD participants. A region similar in size and location was revealed in 45/51 NVA-FTD participants (Figure 3). No brain regions positively correlated to the top 1% of atrophied voxels were detected using these thresholds. These findings were reproduced across a range of atrophy thresholds and individual lesion network map thresholds t ≥ |6|, t ≥ |8| (eFigure 4,5). The occipital cluster was more prominent in VAC-FTD vs. NVA-FTD at the same threshold, but a voxel-wise two sample t-test comparing unthresholded t-maps from VAC-FTD and NVA-FTD found no group differences.

**Figure 3.**
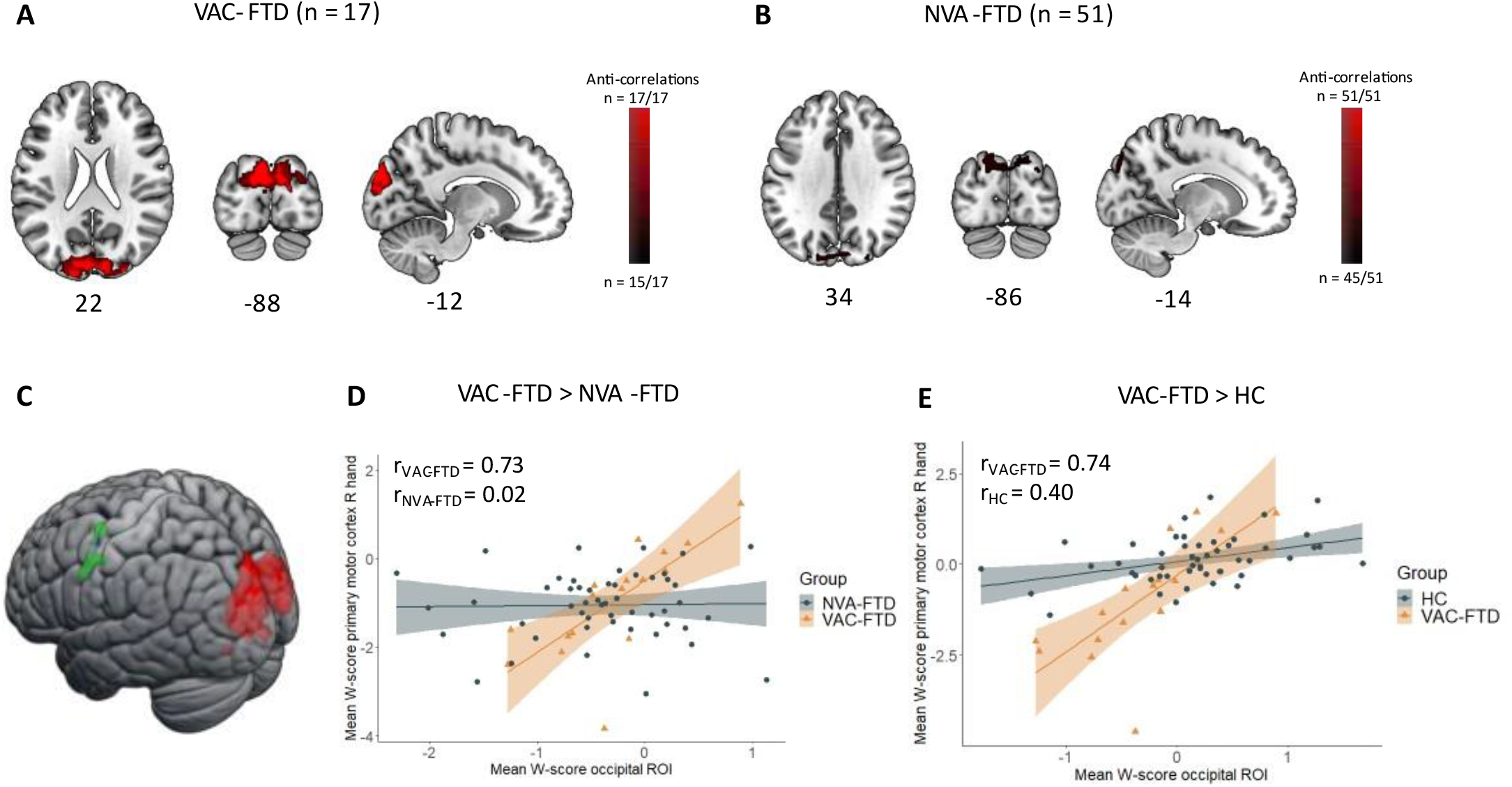
Atrophy network mapping and structural covariance analyses results. **A-B**, Individual atrophy network maps were thresholded at t ≥ |7|, binarized, and overlaid to create group-level atrophy network maps. A similar bilateral dorsomedial occipital region showed anticorrelated brain activity, in healthy controls, to the activity seen in the top 1% of atrophied voxels in both VAC-FTD (n = 17/17 patients) and NVA-FTD (n = 45/51). **C-E**, An interaction model, designed to detect brain regions showing greater structural covariance with the dorsomedial occipital region (red) in VAC-FTD vs. NVA-FTD, identified a cluster in the right hand region of the primary motor cortex (green). This cluster was positively correlated to the mean W-score in the occipital ROI in VAC-FTD but showed no correlation in NVA-FTD (D). A similar cluster (blue, panel C) was observed in a second interaction model aimed to identify regions with greater positive correlation in VAC-FTD than in HC (data shown in E). Findings in C are superimposed on slice and render images of the Montreal Neurological Institute template brain. Images are in neurological orientation (left = left). In D-E, the shaded areas represent the 95% confidence intervals for the fitted regression lines. Abbreviations: VAC-FTD, patients with a frontotemporal dementia spectrum disease and emergence of visual artistic creativity; NVA-FTD, patients with a frontotemporal dementia spectrum disease without emergence of visual artistic creativity; HC, healthy controls; ROI, region of interest; R, right.

### Structural covariance analysis

Having identified a dorsomedial occipital region that anticorrelated with the regions atrophied in FTD, we hypothesized that this region would show unique structural covariation patterns in VAC-FTD vs. NVA-FTD. Structural covariance mapping interrogates neural systems by leveraging between-subject correlations in gray matter volume across the brain ^35,36^. To test our hypothesis, we used the dorsomedial occipital ROI to investigate group differences in inter-regional structural covariance, which may result from long-standing large-scale functional coupling alterations. Our interaction model revealed two clusters significantly correlated with the ROI in VAC-FTD but not in NVA-FTD. The first cluster localized to the left primary motor cortex, in the neighborhood of the motor representation of the right hand (−54, -2, z = 39, k = 437 voxels, cluster level *P* value = 0.012). A second significant cluster was detected in the superior temporal gyrus (58, -39, z = 15, k = 361 voxels, cluster level *P*=0.021). No region showed greater structural covariance in NVA-FTD than in VAC-FTD, and no region showed greater gray matter anticorrelation with the seed in either contrast. We then addressed whether these structural associations were also greater in VAC-FTD than in HC. The second interaction model identified an overlapping cluster in the primary motor cortex (−53, -3, z = 41, k = 183 voxels, cluster level *P* value = 0.045) region but not in the superior temporal gyrus (eTable 5).

### Single case with longitudinal FDG-PET

The cross-sectional analyses presented above suggest the hypothesis that dorsal occipital structure or function becomes enhanced early in FTD, sometimes in association with the appearance of VAC. Remarkably, one patient in the VAC-FTD group, who had a clinical diagnosis of svPPA, underwent FDG-PET scanning fourteen months apart, before and after she started painting. This patient was considered artistically gifted as a child but did not pursue painting until after word finding difficulties emerged. She painted with colored pencils, preferring bright colors and non-human subjects. Leveraging this rare opportunity, we assessed changes in glucose metabolism around the time her creativity blossomed. FDG-PET images obtained before and after VAC onset showed metabolic decline in anterior temporal and frontal regions, as expected, but preserved or increased metabolism in numerous posterior regions (Figure 4 A-B). To quantify these changes across the brain, for each region mean SUVR W-score from the first scan was subtracted from the second scan to produce a regional change map (Figure 4 C-D). Ten brain regions, most in the occipital lobes, showed increased metabolism by more than 0.5 W-score units (eTable 6). Mean regional SUVR W-scores of the dorsomedial occipital ROI uncovered by atrophy network mapping revealed an increase in FDG-PET W-score in parallel to emergence of VAC (1^st^ scan: W = 0.00, 2^nd^ scan: W = 0.83).

**Figure 4.**
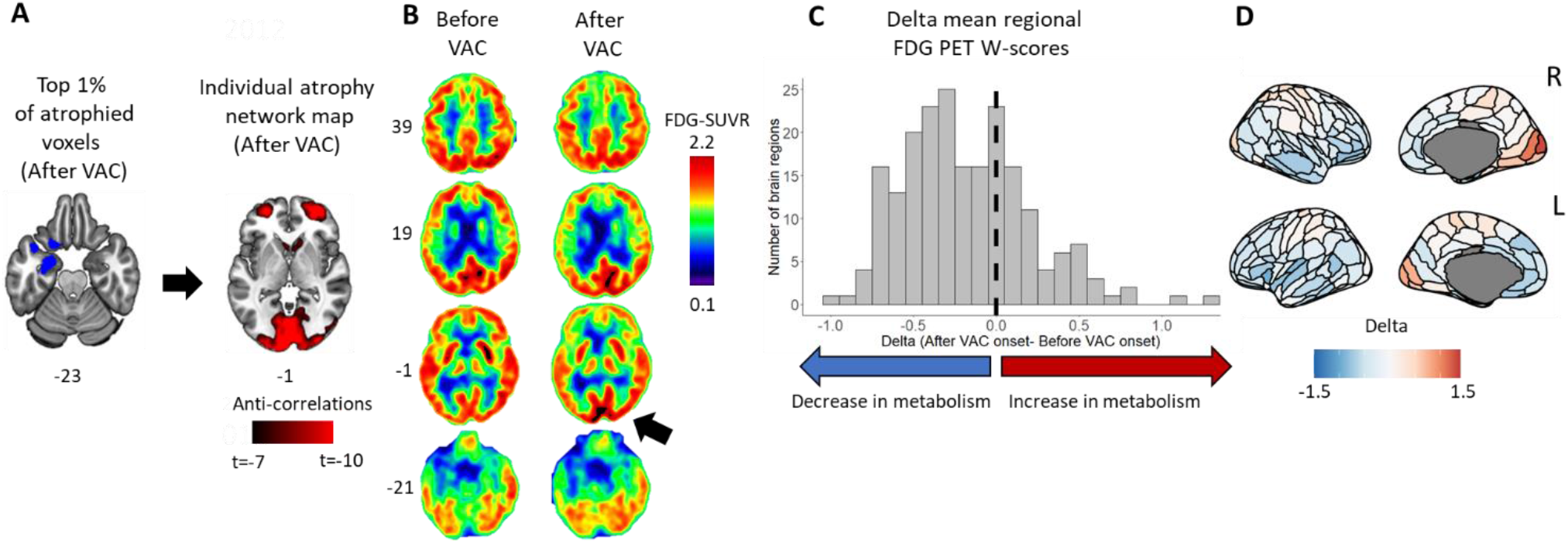
Atrophy network mapping and intensified occipital glucose metabolism after emergence of visual artistic creativity in a patient with svPPA. **A**, Top 1% most atrophied voxels from a patient with svPPA and emergent visual artistic creativity were used as seed to obtain an individual atrophy network map based on the healthy brain connectome. The left anterior temporal lobe regions atrophied in the patient (blue) showed anticorrelated brain activity, in controls, to medial occipital regions (red). **B**, FDG-PET SUVR maps obtained before and after emergence of VAC show declining in glucose metabolism in temporal and frontal regions bilaterally, accompanied by increasing metabolism in medial occipital regions, right greater than left (black arrow). **C,D** Mean regional FDG-PET W-scores were computed for individual brain regions across the whole brain. For each region the W-score after VAC onset was subtracted from that obtained prior to VAC onset to generate a corresponding change value for each region. Multiple brain regions, most prominently primary visual and visual association areas and sensorimotor regions, showed increased metabolism after VAC onset, as mapped on a template brain in (D). Images are in neurological view (left = left). Abbreviations: VAC, visual artistic creativity; FDG-PET, [18F] fluorodeoxyglucose positron emission tomography; SUVR, standardized uptake value ratio, R, right, L left.

## Discussion

The emergence of VAC in FTD occurs in 2.5% of patients and is disproportionately, associated with svPPA. Remarkably, this VAC occurs in the setting of neurodegeneration and results in distinct forms of visual artistic expression. VAC emerges early in the disease course, around the time of FTD symptom onset, as supported by a recent review of single cases reported in the literature^10^.

Atrophy network mapping enables researchers to pinpoint network nodes commonly connected in the healthy brain to lesions from a neurodegenerative disease group of interest. The method is well-suited to uncovering the network basis for aberrant gains of function^31-33^, such as the emergence of VAC studied here. We found that the varied regions of peak frontotemporal atrophy across patients were united by a functional activity pattern that inversely correlated with dorsomedial occipital cortex. This region, encompassing visual association areas V2 and V3 bilaterally, is part of the dorsal visual stream that projects to the posterior parietal cortex^41^. Dorsal stream activity is associated with reaching and grasping behaviors guided by representations of the position, shape, and orientation of objects. Moreover these visual association areas play a pivotal role in representing and relating different aspects of visual images^42^. The inverse functional correlation between FTD atrophy and the dorsal occipital cortex suggests that FTD induces disinhibition of dorsal stream regions which, in turn, predispose some patients to engage in visual art early in the illness. Because only a minority of patients with FTD report intensified VAC, this network rebalancing may manifest as VAC only when certain conditions, such as a latent visual artistic talent or a conducive environment, are met. A role for dorsal occipital regions in driving VAC in FTD received further support from our structural covariance analysis, a between-subjects network mapping technique that can reveal regions that subserve particular behavioral or cognitive functions^43^. Remarkably, VAC-FTD patients, when compared to those with NVA-FTD, demonstrated greater structural covariance between dorsomedial occipital cortex and the left primary motor cortex, around the representation of the right hand. In our view, this effect likely reflects the visuomotor activities that emerge when dorsomedial occipital cortex hyperactivity drives visual creativity in an artistically predisposed brain operating within a conducive environment. In this context, plastic cortical remodelling may enhance the structural correlation between visual and motor areas, reflecting patient’s new preoccupation^44,45^. Our PET-based single case with increasing occipital glucose metabolism in parallel to the emergence of VAC also supports the notion that hyperactivation of the dorsal stream predisposes to visual artistic engagement.

This work provides convergent structural and functional evidence that FTD lesion-induced intensification of dorsal visual association areas predisposes to the emergence of VAC, a gain of function that emerges early in the illness. One limitation of this study is the small VAC-FTD sample. Although it is by far the largest VAC-FTD cohort reported to date, the sample size may have precluded detection of additional VAC-relevant brain regions. Future longitudinal studies may clarify not only the emergence of VAC in FTD but also the onset of other enhanced capacities early in the course of aging and neurodegeneration.

## Supporting information

Supplementary Online Content

## Data Availability

All data produced in the present study are available upon reasonable request to the authors

