## Supplementary Online Content for "Emergent visual creativity in frontotemporal dementia is associated with dorsomedial visual cortex enhancement"

Friedberg A, Pasquini L et al.

eMethods 1: Clinical and neuropsychological measures
eMethods 2: MRI quality control
eMethods 3: Creation of individual W-score maps (individual atrophy maps)
eMethods 4: Functional MRI data preprocessing
eMethods 5: $^{18}$F-labelled fluorodeoxyglucose positron emission tomography (FDG-PET) images acquisition and preprocessing
eMethods 6: Generation of FDG-PET W-score maps
eTable 1. Structural T1-weighted image acquisition protocols by scanner.
eTable 2. Demographic characteristics of patient groups and 175 healthy controls used to derive individual atrophy network maps
eTable 3. Characteristics of patients with emergence of visual artistic creativity and frontotemporal dementia
eTable 4. Neuropathological diagnoses of patients with frontotemporal dementia and emergence of visual artistic creativity (n = 6/17)
eTable 5. Interaction models results
eTable 6. Brain regions that increased in FDG-PET metabolism more than 0.5 W-scores in the patient scanned before and after onset of visual artistic creativity
eFigure 1: Atrophy network mapping technique.
eFigure 2: Mean W-score maps: VAC-FTD, NVA-FTD
eFigure 3: Frequency maps: VAC-FTD, NVA-FTD
eFigure 4: Atrophy network mapping control analyses – controlling for seed size
eFigure 5: Atrophy network mapping control analyses – different t thresholds

### Supplementary methods

#### eMethods 1: Clinical and neuropsychological measures

All participants provided written informed consent, and the University of California, San Francisco, Committee on Human Research approved the study. All neurological and neuropsychological assessments occurred within 180 days of MRI scanning that was used as index. The Neuropsychiatric Inventory Questionnaire (NPI-Q) was used to examine differences in neuropsychiatric behavioral alterations between VAC-FTD and NVA-FTD (table 1). The NPI-Q rates the existence, frequency, and severity of neuropsychiatric behavioral alterations within the last month across twelve domains[1]. Information with regards to frequency of abnormal behaviors across the twelve domains measured by the NPI-Q was not systematically collected in both VAC-FTD and NVA-FTD groups. We used the information with regards to the presence (YES/NO) of each abnormal behavior in between-group comparisons which was available for all participants and was less influenced by informant's characteristics (gender, cultural background etc.). Between-group differences in clinical and neuropsychological characteristics were assessed using t-test, Mann-Whitney test, analysis of variance (ANOVA), or Kruskal-Wallis test for continuous variables and chi-square and Fisher's exact test for categorical data, as appropriate. Test statistics were considered significant at $P < 0.05$ (two-tailed). Statistical analyses were performed with R, version 4.1.1 (R Foundation for Statistical Computing).

#### eMethods 2: MRI quality control

All MRI images were visually inspected by AF, IIG or LP. Additionally, we obtained MRI quality control measures using the CAT12 toolbox in SPM12 (running in MATLAB r2018b). Following these procedures all images with significant motion artifacts or lesions were excluded from further analyses.

#### eMethods 3: Creation of individual W-score maps (individual atrophy maps)

To generate the W-score model[2,3], we first performed a voxel-wise general linear model (GLM) for the smoothed images of a cohort of 397 cognitively normal older adults assessed at the UCSF Memory and Aging Center. Age at MRI, sex, handedness, years of education (YOE), scanner type (three distinct covariates, one for each scanner type) and total intracranial volume (TIV) were included as covariates. This sample had the following demographic characteristics: Mean age at MRI (standard deviation (SD)) = 69.3 (8.8) years; mean YOE (SD) = 17.2 (2.3) years; mean TIV (SD) = 1.4 (0.1) liters; male/female = 158/239; handedness right/left = 354/43; scanner 1.5T/3T/Prisma/4T = 58/144/140/52. While most demographical parameters were evenly distributed across sex and scanner type, an ANOVA model revealed significant differences in age across scanner type ($F = 12.7$; $p<0.001$). The smoothed images were subsequently used to derive an initial W-score model that included interaction terms for age and scanner type:

$$GM_{CON} = \beta 0 + \beta 1 * Age + \beta 2 * Sex + \beta 3 * Handedness + \beta 4 * TIV + \beta 5 * Scanner\_1 + \beta 6 * Scanner\_2 + \beta 7 * Scanner\_3 + \beta 9 * YOE + \beta 10 * Scanner\_1 * Age + \beta 11 * Scanner\_2 * Age + \beta 12 * Scanner\_3 * Age + \varepsilon$$

where $GM_{CON}$ is the voxel-specific value of a segmented gray matter tissue density map in the control sample.
To estimate the statistical significance of each parameter coefficient map, corresponding t-maps were derived by dividing the parameter coefficient maps by the standard deviation of the residuals. These analyses revealed a negligible effect of scanner type*age interaction on expected gray matter intensity in our sample. Therefore, a final W-score model was estimated without the scanner type*age interaction terms:

$$GM_{CON} = \beta 0 + \beta 1 * Age + \beta 2 * Sex + \beta 3 * Handedness + \beta 4 * TIV + \beta 5 * Scanner\_1 + \beta 6 * Scanner\_2 + \beta 7 * Scanner\_3 + \beta 8 * YOE + \varepsilon$$

Subsequently, individual W-score maps were computed for the patients as follows:

$$W = \frac{Observed - Expected}{SD_{\varepsilon}}$$

*Observed* is the raw value of a voxel from the smoothed image of a patient; *Expected* is the expected value for the voxel of a specific patient adjusted for covariates using the healthy control model; and SDε is the standard deviation of the residuals from the healthy control model.

### eMethods 4: Functional MRI data preprocessing

Functional MRI scans were processed using fMRIPrep[4] (RRID:SCR_016216). For anatomical image processing, the MPRAGE images were corrected for intensity non-uniformity (INU) with N4BiasFieldCorrection in ANTs [5], 2008) (RRID:SCR_004757), and used as T1w-reference throughout the workflow. The T1w-reference was skull-stripped with a Nipype[6] (RRID:SCR_002502) implementation of the antsBrainExtraction.sh workflow using OASIS30ANTs as target template. Brain tissue segmentation of cerebrospinal fluid (CSF), white-matter (WM) and gray-matter (GM) was performed on the brain-extracted T1w using FSL fast (https://fsl.fmrib.ox.ac.uk/fsl/fslwiki; RRID:SCR_002823). Volume-based spatial normalization to the MNI152NLin6Asym standard space was performed through nonlinear registration with antsRegistration, using brain-extracted versions of both T1w reference and the T1w template.

For functional image processing, the first five volumes were removed to allow for scanner equilibration. A reference volume and its skull-stripped version were generated by fMRIPrep. The BOLD reference was then co-registered to the T1w reference using FSL flirt with 6-degrees-of-freedom affine registration. Co-registration was configured with nine degrees of freedom to account for distortions remaining in the BOLD reference. Head-motion parameters with respect to the BOLD reference (transformation matrices, and six corresponding rotation and translation parameters) were estimated using FSL mcflirt and were used to compute the framewise displacement (FD). BOLD runs were slice-time corrected using AFNI 3dTshift (https://afni.nimh.nih.gov/; RRID:SCR_005927). The BOLD images were realigned from native to MNI152NLin6Asym standard space using antsApplyTransforms, configured with Lanczos interpolation, with a single interpolation step by composing transformations for head-motion and co-registrations to anatomical and output spaces. Images were spatially smoothed with a 6mm FWHM (full-width half-maximum) kernel using FSL susan. Confounding CSF and WM timeseries were calculated based on the preprocessed BOLD images, deriving average signals using the subject-specific anatomically derived tissue masks after erosion. The confound timeseries for head motion estimates, CSF, and WM were expanded with the inclusion of temporal derivatives and quadratic terms for each[7]. Bandpass filtering in the frequency range 0.008-0.08 Hz was performed on the confound timeseries and BOLD images using fslmaths and AFNI 3dBandpass respectively. Confound timeseries were then regressed out of the BOLD images using fslglm. Subjects with greater than 0.55 mm mean FD were excluded from subsequent analysis[8].

### eMethods 5: $^{18}$F-labelled fluorodeoxyglucose positron emission tomography (FDG-PET) images acquisition and preprocessing

#### PET radiochemistry and acquisition

[18F]FDG was purchased from a commercial vendor (IBA Molecular). PET scans were performed at Lawrence Berkeley National Laboratory using a Siemens ECAT EXACT HR PET scanner in 3-dimensional acquisition mode. 30 minutes of dynamic FDG data were obtained. Ten-minute transmission scans for attenuation correction were obtained either immediately before or after each[18F]FDG scan. PET data were reconstructed using an ordered subset expectation maximization algorithm with weighted attenuation. Images were smoothed with a 4mm Gaussian kernel with scatter correction. All images were evaluated before analysis for patient motion and adequacy of statistical counts.

#### PET pre-processing and analysis

All image processing and analysis was performed in Statistical Parametric Mapping version 12 (SPM12; http://www.fil.ion.ucl.ac.uk/spm). Reference regions were defined in native MRI space for each subject using subcortical parcellations from FreeSurfer. FDG-PET frames were summed and standard uptake volume ratios (SUVR) were calculated by normalizing the summed FDG image to mean activity in the pons for each subject[9].

#### Spatial normalization

FDG data was co-registered to the subject's skull stripped T1-weighted MRI. To allow across-subject comparisons, each subject's T1-weighted MRI was normalized to MNI (Montreal Neurological Institute) space using the skull stripped ch2 template, and the derived normalization parameters were applied to the subject's co-registered FDG volumes. All normalized images were smoothed with a 12-mm Gaussian kernel.

**Partial volume correction**

In a post-hoc analysis, a two-compartmental partial-volume correction to all MRI scans was applied in order to correct PET data for atrophy [10]. The correction procedure involved convolving a binary brain mask (a sum of grey and white matter segmented images from the subject's T1-weighted MRI obtained from FreeSurfer, eroded by one voxel) with the point-spread function specific to the PET tomography along all axes. This provided a means for estimating the percentage of brain tissue emitting radioactivity at each voxel. The PET counts for each voxel were then adjusted based on the percentage of estimated brain matter[11].

### eMethods 6: Generation of FDG-PET W-score maps

For these statistical maps, we used a W-score model generated based on FDG PET scans derived from 71 healthy controls who were included in the Berkeley Aging Cohort Study (BACS) and Neuroimaging in Frontotemporal Dementia (NIFD) study. Details on BACS inclusion criteria can be found in previous publications[12]. For up-to-date information on NIFD participation and protocol, please visit http://memory.ucsf.edu/research/studies/nifd. Demographic characteristics of 71 controls used to generate the model were the following: mean age at scan (SD) = 68(15), male/female = 32/39, handedness right/left = 67/4, mean years of education (SD) = 16.8(2), mean MMSE(SD) = 28.9 (1.1). In this model W-scores were adjusted for age, sex handedness and years of education.

### eTable 1. Structural T1-weighted image acquisition protocols by scanner.

| | Scanner I | Scanner II | Scanner III | Scanner IV |
|---|---|---|---|---|
| **Manufacturer (system)** | Siemens (Tim Trio) | Siemens (Magnetom Prisma) | Siemens (Magnetom VISION) | Siemens (Bruker MedSpec) |
| **Magnet strength** | 3T | 3T | 1.5T | 4T |
| **Repetition time (ms)** | 2300 | 2500 | 5000 | 2300 |
| **Echo time (ms)** | 2.98 | 2.82 | 20 | 3 |
| **Slice Thickness (mm)** | 1 | 1 | 1.5 | 1 |
| **Voxel size (mm)** | 1 x 1 x 1 | 1 x 1 x 1 | 1.5 x 1.5 x 1.5 | 1 x 1 x 1 |
| **Groups of scanned participants, VAC-FTD/ NVA-FTD/HC/HC-ATN** | 3/11/51/175 | 5/24/0/0 | 7/13/0/0 | 2/3/0/0 |

Abbreviations: mm = millimeters; ms = milliseconds; VAC-FTD = patients with emergence of visual artistic creativity and frontotemporal dementia spectrum diseases, NVA-FTD = patients without emergence of visual artistic creativity and frontotemporal dementia spectrum diseases, HC=healthy controls, HC-ATN = healthy controls that were used for generation of individual atrophy network maps.

### eTable 2. Demographic characteristics of patient groups and 175 healthy controls used to derive individual atrophy network maps

| | VAC-FTD | NVA-FTD | HC-ATN | ***P*-value** |
|---|---|---|---|---|
| n | 17 | 51 | 175 | |
| Age at MRI[a], years | 65(9.7) | 64.8(7) | 65.0(8.6) | 0.99 |
| Male:Female[b], n | 7:10 | 26:25 | 85:90 | 0.78 |
| Right:Left[b], n | 14:3 | 42:9 | 159:16 | 0.18 |
| Education[c], years | 16.1(4.7) | 16.6(2.1) | 17(2.2) | 0.44 |

[a] One way analysis of variance (ANOVA)
[b] Chi square test
[c] Kruskal-Wallis test
Abbreviations: VAC-FTD = patients with emergence of visual artistic creativity and frontotemporal dementia spectrum diseases, NVA-FTD = patients without emergence of visual artistic creativity and frontotemporal dementia spectrum diseases, HC-ATN = healthy controls that were used for generation of individual atrophy network maps

### eTable 3. Characteristics of patients with emergence of visual artistic creativity and frontotemporal dementia

| Patient no. | Age at first FTD symptom | Age artistic skills emerge | Delta | Clinical diagnosis | Change in visual artistic creativity | Primary modality of visual art | Past artistic interest[a] |
|---|---|---|---|---|---|---|---|
| 1 | 51-55 | 56-60 | 5 | svPPA | Increase in quantity, change in style of visual art | Painting | ++ |
| 2 | 41-45 | 56-60 | 16 | bvFTD | De novo emergence of visual artistic creativity | Sculpting from tin objects | - |
| 3 | 56-60 | 61-65 | 1 | svPPA | De novo emergence of visual artistic creativity | Sculpture, Installation art | - |
| 4 | 66-70 | 66-70 | 1 | nfvPPA | De novo emergence of visual artistic creativity | Painting, making jewelry, glass painting | - |
| 5 | 46-50 | 46-50 | 0 | svPPA | Change in style | Painting | ++ |
| 6 | 46-50 | 46-50 | -1 | svPPA | De novo emergence of visual artistic creativity | Painting, sculpture | - |
| 7 | 51-55 | 51-55 | 3 | svPPA | Change of modality from dance and music to verbal and visual art | Photography | + |
| 8 | 41-45 | 46-50 | 2 | bvFTD | De novo emergence of visual artistic creativity | Color pencil illustrations | - |
| 9 | 46-50 | 56-60 | 9 | bvFTD | Recrudescence of interest in visual art from his twenties | Painting, photography | + |
| 10 | 61-65 | 31-35 | -30 | svPPA | Change of modality from dancing and opera singing to visual art | Pottery | + |
| 11 | 56-60 | 61-65 | 3 | PSP-RS | De novo emergence of visual artistic creativity | Sculpture in clay | - |
| 12 | 56-60 | 41-45 | -16 | PSP-RS | Recrudescence of interest in visual art from her twenties | Quilting | + |
| 13 | 66-70 | 71-75 | 3 | svPPA | Reemergence of dormant artistic talent from childhood | Painting | + |
| 14 | 56-60 | 51-55 | -8 | nfvPPA | Increase in quantity, change in style | Painting | + |
| 15 | 81-85 | 86-90 | 1 | CBS | Increase in quantity | Painting | + |
| 16 | 56-60 | 56-60 | 2 | svPPA | De novo emergence of visual artistic creativity | Creating montages with superimposed poems | - |
| 17 | 56-60 | 56-60 | -3 | ALS | De novo emergence of visual artistic creativity | Making jewelry | + |

[a] Past artistic interest: (++) Professional artist who experienced change in style (+) Visual art was a prior minor hobby and/or prior visual artistic education was obtained and /or there was any past interest in nonvisual art (-) No prior past artistic interest.

Abbreviations: svPPA = semantic variant of primary progressive aphasia, bvFTD = behavioral variant of frontotempotral dementia, nfvPPA = nonfluent variant of primary progressive aphasia, PSP-RS = progressive supranuclear palsy - Richardson syndrome, CBS, corticobasal syndrome, ALS amyotrophic lateral sclerosis.

### eTable 4. Neuropathological diagnoses of patients with frontotemporal dementia and emergence of visual artistic creativity (n = 6/17)

| Patient no.[a] | Primary neuropathological diagnosis | Secondary neuropathological diagnosis | ADNC level | LBD Stage |
|---|---|---|---|---|
| 2 | FTLD-TDP, Type B | Motor neuron disease, lower motor neuron only | Not ADNC | None |
| 3 | FTLD-tau, Pick's disease | | Low | None |
| 5 | FTLD-TDP, Type C | | Low | None |
| 6 | FTLD-TDP, Type A | Amyotrophic lateral sclerosis | Not to low | None |
| 12 | FTLD-tau, Corticobasal degeneration | | Not ADNC | None |
| 14 | FTLD-tau, Corticobasal degeneration | | Not to low | None |

[a] Patient no. as in eTable 3. Abbreviations: FTLD=frontotemporal degeneration, ADNC=Alzheimer’s disease neuropathological change, LBD= Lewy body disease, TDP= transactive response DNA-binding protein of 43 kDa

### eTable 5. Interaction models results

| **Model** | | | **x,y,z** | **T** | **Z** | **Region** | **BA** |
|---|---|---|---|---|---|---|---|
| Interaction model VAC-FTD>NVA-FTD, MMSE as covariate of no interest | | | | | | | |
| | Cluster 1 (437 voxels) | | | | | | |
| | | Maximum 1 | -54,-2,39 | 4.68 | 4.32 | L premotor cortex | 6 |
| | | Maximum 2 | -52,-10,46 | 4.02 | 3.78 | L primary motor cortex | 4 |
| | | Maximum 3 | -39,-3,60 | 4.01 | 3.77 | L premotor cortex | 6 |
| | Cluster 2 (361 voxels) | | | | | | |
| | | Maximum 1 | -58,-39,15 | 4.49 | 4.17 | L superior temporal gyrus | 22 |
| | | Maximum 2 | -54,-30,9 | 3.7 | 3.51 | L primary auditory cortex | 41 |
| | | Maximum 3 | -62,-24,10 | 3.67 | 3.48 | L primary auditory cortex | 41 |
| Interaction model VAC-FTD>NVA-FTD | | | | | | | |
| | Cluster 1 (282 voxels) | | | | | | |
| | | Maximum 1 | -54,-2,39 | 4.43 | 4.12 | L premotor cortex | 6 |
| Interaction model VAC-FTD>HC | | | | | | | |
| | Cluster 1 (183 voxels) | | | | | | |
| | | Maximum 1 | -53,-3,41 | 4.96 | 4.54 | L premotor cortex | 6 |
| | | Maximum 2 | -41,2,50 | 3.88 | 3.67 | L premotor cortex | 6 |
| | Cluster 2 (285 voxels) | | | | | | |
| | | Maximum 1 | -45,29,26 | 4.86 | 4.46 | L DLPFC | 9 |
| | | Maximum 2 | -45,24,33 | 4.8 | 4.42 | L DLPFC | 10 |
| | | Maximum 3 | -41,11,32 | 3.87 | 3.66 | L frontal eye field | 8 |

| | | | | | | | |
|---|---|---|---|---|---|---|---|
| | Cluster 3 (443 voxels) | | | | | | |
| | | Maximum 1 | 41,14,48 | 4.69 | 4.33 | R Broca | 44 |
| | | Maximum 2 | 45,15,33 | 4.13 | 3.87 | R Broca | 45 |
| | | Maximum 3 | 45,2,50 | 3.73 | 3.53 | R premotor cortex | 6 |
| | Cluster 4 (290 voxels) | | | | | | |
| | | Maximum 1 | 18,35,48 | 4.68 | 4.32 | R frontal eye field | 8 |
| | | Maximum 2 | 14,44,41 | 4 | 3.76 | R DLPFC | 9 |
| | | Maximum 3 | 6,44,33 | 3.45 | 3.29 | R dorsal ACC | 32 |
| | Cluster 5 (201 voxels) | | | | | | |
| | | Maximum 1 | -47,-11,-39 | 4.54 | 4.21 | L primary motor cortex | 4 |
| | | Maximum 2 | -53,-5,-24 | 3.73 | 3.54 | L medial temporal gyrus | 21 |
| | | Maximum 3 | -53,-2,-33 | 3.7 | 3.51 | L temporal pole | 38 |
| | Cluster 6 (326 voxels) | | | | | | |
| | | Maximum 1 | 62,-41,18 | 4.38 | 4.08 | R superior temporal gyrus | 22 |
| | | Maximum 2 | 56,-36,44 | 4.09 | 3.84 | R supramarginal gyrus | 40 |
| | | Maximum 3 | 57,-41,36 | 3.46 | 3.3 | R supramarginal gyrus | 41 |
| | Cluster 7 (224 voxels) | | | | | | |
| | | Maximum 1 | -30,-51,-51 | 4.23 | 3.95 | L cerebellar hemisphere | NA |
| | Cluster 8 (189 voxels) | | | | | | |
| | | Maximum 1 | 44,-2,-41 | 3.98 | 3.75 | R inferior temporal gyrus | 20 |
| | | Maximum 2 | 48,-12,-38 | 3.85 | 3.64 | R inferior temporal gyrus | 21 |
| | | Maximum 3 | 48,5,-33 | 3.52 | 3.35 | R temporal pole | 38 |
| | Cluster 9 (180 voxels) | | | | | | |

| | | | | | | | |
|---|---|---|---|---|---|---|---|
| | | Maximum 1 | -50,24,9 | 3.98 | 3.75 | L Broca area | 45 |
| | | Maximum 2 | -53,15,15 | 3.93 | 3.7 | L Broca area | 44 |

Abbreviations: VAC-FTD = patients with emergence of visual artistic creativity and frontotemporal dementia spectrum diseases, NVA-FTD = patients without emergence of visual artistic creativity and frontotemporal dementia spectrum diseases, HC = healthy controls, MMSE =mini mental status examination, BA= Broadmann area, R= right, L=left.

**eTable 6: Brain regions that increased in FDG-PET metabolism more than 0.5 W-scores in the patient scanned before and after onset of visual artistic creativity**

| Brainnetome region number | Right/Left | Lobe | Gyrus | Delta |
|---|---|---|---|---|
| 106 | Right | Temporal lobe | Fusiform Gyrus | 0.64 |
| 117 | Left | Temporal lobe | Parahippocampal Gyrus | 0.53 |
| 189 | Left | Occipital Lobe | Medioventral Occipital Cortex | 0.61 |
| 190 | Right | Occipital Lobe | Medioventral Occipital Cortex | 0.67 |
| 191 | Left | Occipital Lobe | Medioventral Occipital Cortex | 0.77 |
| 192 | Right | Occipital Lobe | Medioventral Occipital Cortex | 1.08 |
| 193 | Left | Occipital Lobe | Medioventral Occipital Cortex | 0.82 |
| 194 | Right | Occipital Lobe | Medioventral Occipital Cortex | 1.31 |
| 196 | Right | Occipital Lobe | Medioventral Occipital Cortex | 0.65 |
| 200 | Right | Occipital Lobe | Lateral Occipital Cortex | 0.51 |

### eFigure 1: Atrophy network mapping technique.

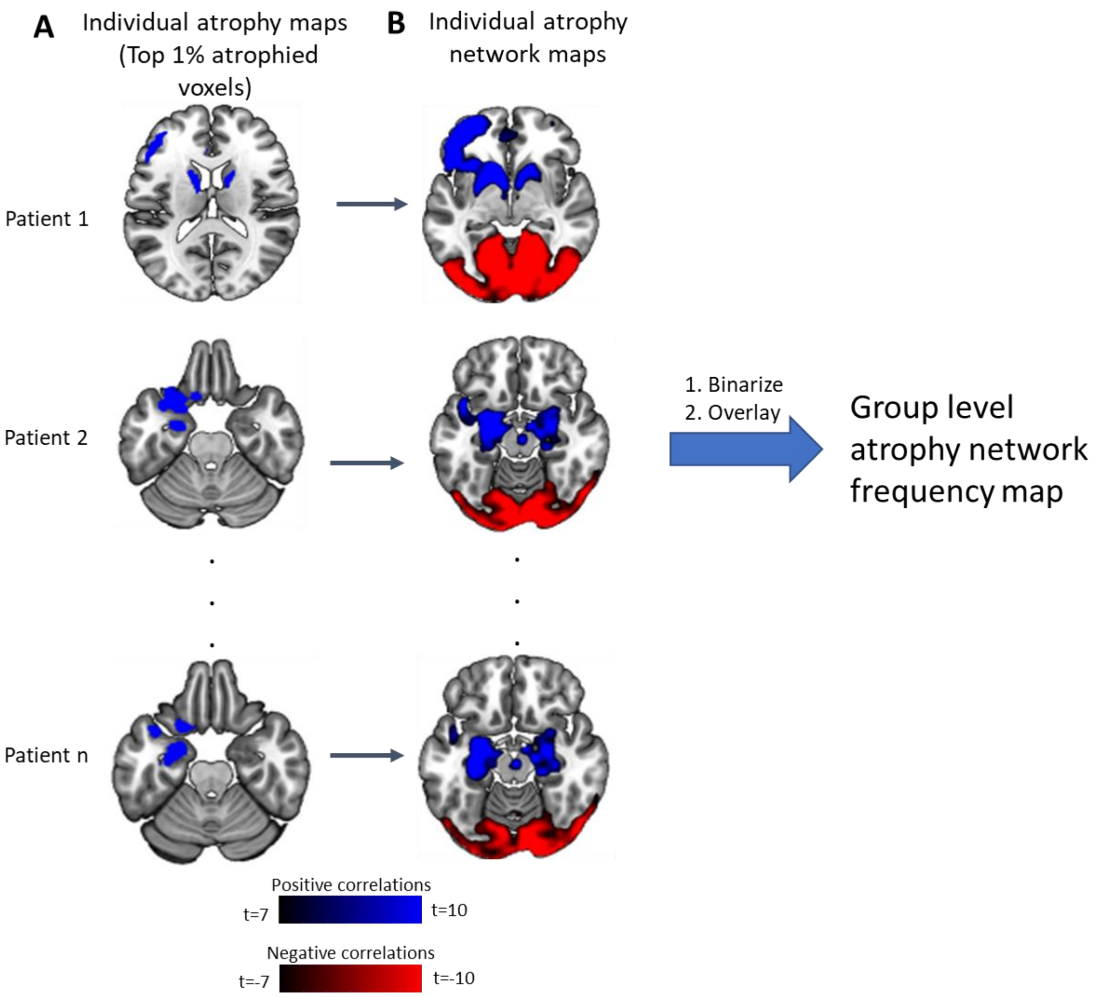

(A) Each patient's W-score map was binarized to generate individual atrophy maps (top 1% most atrophied voxels). (B) Regions positively and negatively functionally connected to each patient's atrophy map based on a connectome of matched healthy controls (n=175). Individual atrophy network maps of each group were thresholded, binarized and overlayed to generate group level atrophy network frequency maps.

### eFigure 2: Mean W-score maps: VAC-FTD, NVA-FTD

**VAC-FTD (-5<W<-2)**

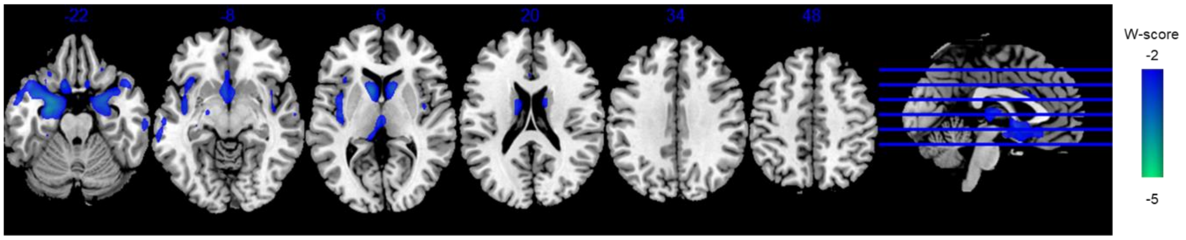

**NVA-FTD (-5<W<-2)**

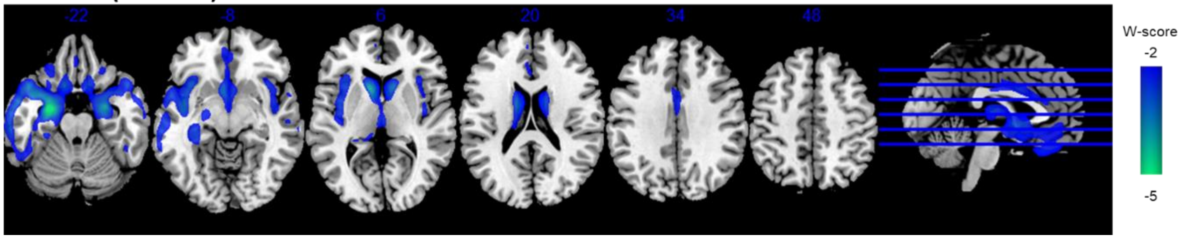

Findings are projected on Montreal Neurological Institute template brain. Images are in neurological orientation (left = left).
Abbreviations: VAC-FTD = patients with emergence of visual artistic creativity and frontotemporal dementia spectrum diseases, NVA-FTD = patients without emergence of visual artistic creativity and frontotemporal dementia spectrum diseases.

### eFigure 3: Frequency maps: VAC-FTD, NVA-FTD

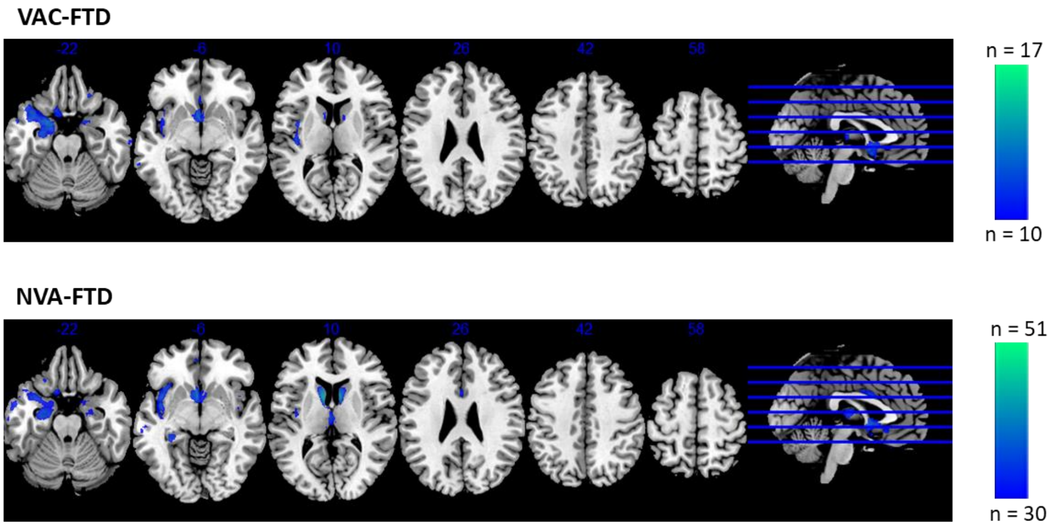

Individual W-score maps were thresholded at W<-2, binarized and overlayed to produce frequency maps for each group. No clusters were demonstrated above n=12/17 VAC-FTD patients. Findings are projected on Montreal Neurological Institute template brain. Images are in neurological orientation (left = left). Abbreviations: VAC-FTD = patients with emergence of visual artistic creativity and frontotemporal dementia spectrum diseases, NVA-FTD = patients without emergence of visual artistic creativity and frontotemporal dementia spectrum diseases.

### eFigure 4 Atrophy network mapping control analyses – controlling for seed size

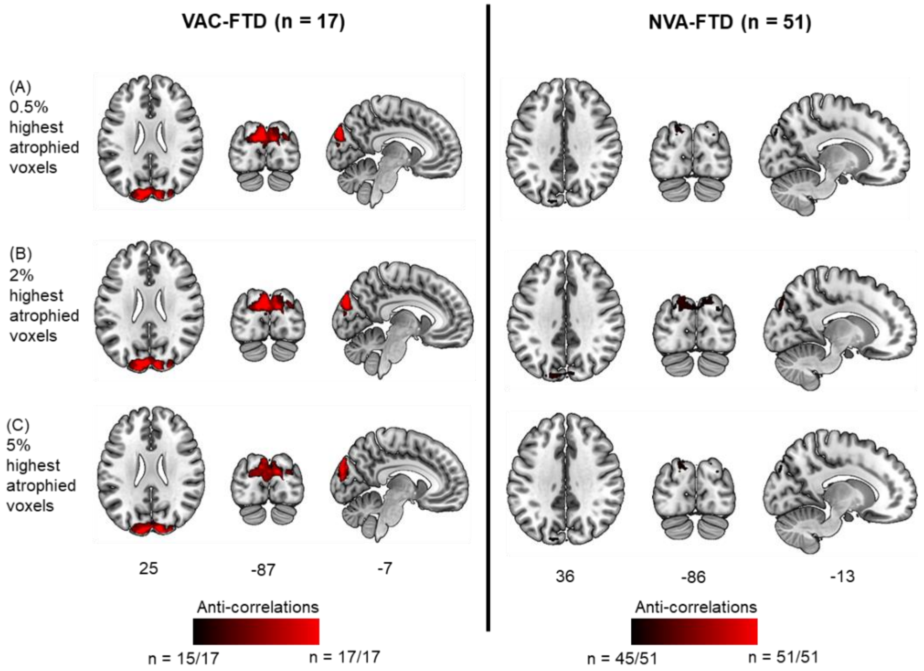

Group level atrophy network maps, generated by using seed regions-of-interest derived from the highest 0.5%,2% and 5% of atrophied voxels from individual patient atrophy maps, resulted in similar findings. Individual atrophy network maps were thresholded and binarized at |t| ≥7 and overlaid to produce group level atrophy network maps. No positive correlations to the atrophy patterns were detected using these thresholds. Findings are projected on Montreal Neurological Institute template brain. Images are in neurological orientation (left = left).

Abbreviations: VAC-FTD = patients with emergence of visual artistic creativity and frontotemporal dementia spectrum diseases, NVA-FTD = patients without emergence of visual artistic creativity and frontotemporal dementia spectrum diseases.

### eFigure 5 Atrophy network mapping control analyses – different t thresholds

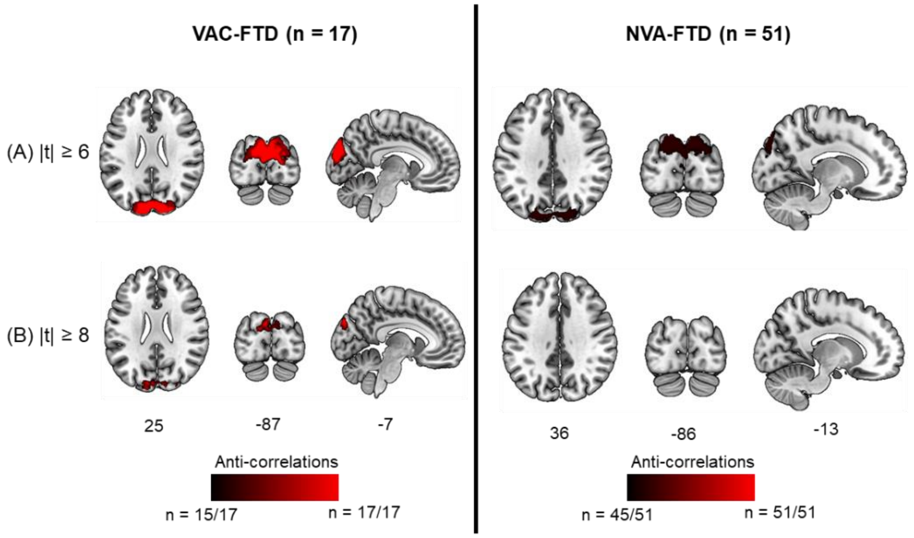

Individual atrophy network maps generated by seeding top 1% of atrophied voxels were thresholded at alternative statistical thresholds (|t|≥6, |t|≥8). Findings were overlaid to produce group level atrophy network frequency maps, which were similar those derived using |t|≥7. No positive correlations to the atrophy patterns were detected. Findings are projected on Montreal Neurological Institute template brain. Images are in neurological orientation (left = left).
Abbreviations:  VAC-FTD = patients with emergence of visual artistic creativity and frontotemporal dementia spectrum diseases, NVA-FTD = patients without emergence of visual artistic creativity and frontotemporal dementia spectrum diseases.
